# Reducing Wear Rate of Precision Surgical Instruments through Lean Six Sigma: A single-center retrospective study

**DOI:** 10.1101/2025.06.02.25328841

**Authors:** Xiaoxia Zhu, Jing Qin, Xia Zhou, HaiPing Chen, CongBing Yan, Rong Bao

## Abstract

**Background:** With the continuous advancement of technology and rapid development of precision medicine, precision medical devices are increasingly being utilized in clinical diagnosis and treatment. These devices play a crucial role in clinical practice. However, improper use accelerates instrument wear and tear, increases hospital costs, and jeopardizes patient safety. In Mainland China, research on surgical instrument wear is limited. Healthcare institutions widely implement Lean Six Sigma (LSS) as a management tool to enhance medical quality and patient safety through process improvement.

**Objective:** This study aimed to reduce the wear rate of precision instruments using the LSS management method, thereby lowering medical costs and enhancing patient safety. It also seeks to provide recommendations to strengthen the quality management of hospitals.

**Methods:** The study applied five LSS phases (Define, Measure, Analyze, Improve, Control) to analyze instrument deterioration. The primary causes of instrument degradation were identified, and three major improvement plans were proposed: establishing an intelligent device traceability and sterile supply chain quality control management system, refining transportation management, and optimizing the instrument preprocessing position. This study compared the wear and tear rates of precision instruments before and after implementation of the LSS method from 2023 to 2024.

**Results:** The wear rate decreased significantly from 17.63% to 6.54% (P<0.001), yielding direct cost savings of 769,000 Chinese Yuan (CNY),calculated from the reduced repair and replacement expenses. Furthermore, the satisfaction rates among surgeons and nurses rose from 83.33% to 95.83% and from 86.67% to 98.33%, respectively (P<0.05).

**Conclusion:** The full lifecycle management of precision instruments based on LSS can effectively reduce the wear rate, lower hospital costs, and improve the satisfaction of surgeons and nurses in sterile supply departments.

**Data Access Statement:** The raw data used in this study are proprietary and cannot be shared publicly due to department confidentiality agreements.

## 1 INTRODUCTION

High-precision medical devices (e.g., orthopedic power systems, endoscopic tools, and neurosurgical microinstruments) are now used in over 50% of clinical procedures, reflecting their critical role in modern healthcare^[1]^. High-precision and sensitive medical devices are critical in clinical practice. These devices detect subtle physiological changes that are often missed by traditional methods, which improves diagnostic accuracy and treatment success rates. However, they can accurately reach the lesion sites within the patient’s body to perform treatments while protecting the surrounding healthy tissues from damage. This not only reduces the incidence of postoperative complications but also accelerates patient recovery, significantly improving the quality of rehabilitation^[2]^.

Despite their significant advantages in clinical practice, precision medical devices still face challenges. First, owing to their complex and delicate structures and susceptibility to damage, managing these devices in a clinical setting is relatively difficult. Improper operation or inadequate maintenance can affect surgical procedures and even cause new trauma to patients, posing potential safety risks^[3 -4]^. Second, given their high cost and limited supply, increased wear and tear directly increase hospital operating costs and complicate resource allocation and patient treatment planning^[5 - 6]^. Therefore, the application of precision medical devices is a double-edged sword, and maximizing their positive impacts while minimizing potential risks is an urgent issue in the current medical field.

Lean Six Sigma is a widely implemented management tool and process improvement technique in the healthcare sector^[7-8]^. Six Sigma is a quality management strategy aimed at improving process quality by identifying and eliminating defects. It employs a structured problem-solving approach described by the acronym “DMAIC,” which stands for Define, Measure, Analyze, Improve, and Control^[9]^. On the other hand, lean focuses on eliminating waste and non-value-added activities to increase speed and reduce operational costs, emphasizing the understanding of value from the customer’s perspective and identifying activities that do not meet customer needs as part of the process to be eliminated^[10]^. Lean Six Sigma (LSS) integrates lean waste reduction principles with Six Sigma’s focus on minimizing process variability, offering a systematic approach to quality improvement ^[8, 11]^.

In the work practice of a comprehensive hospital in China, it was observed that the degradation rate of precision instruments has increased year by year. This increase directly leads to higher hospital costs. Moreover, the wear and tear of precision instruments increases the uncertainty in surgical procedures, posing potential threats to patient health and safety and causing strong dissatisfaction among surgeons and nurses.

We hypothesized that applying lean six-sigma (LSS) methods to the management of precision instruments could reduce the wear rate and hospital costs. Based on this hypothesis, the research team used the LSS method to implement a series of management improvements for the entire lifecycle of precision instruments and conducted a before-and-after cohort study to compare wear and tear rates and the direct medical costs of precision instruments in the hospital.

## 2 METHODS

### 2.1 Design

This was a before-and-after cohort study. The control group comprised instruments processed from January to October 2023 (pre-LSS), whereas the experimental group included those processed during the same period in 2024 (post-LSS). To minimize confounding effects, the hospital maintained consistent staffing levels and operational policies during both the study periods. No additional training programs or equipment maintenance protocols beyond the LSS interventions were introduced in 2024.

### 2.2 Setting

The study was conducted in the Central Sterile Supply Department of the First Affiliated Hospital of Naval Medical University in Shanghai.

### 2.3 Participants

The inclusion criteria were as follows: ➀ Precision instruments were defined as medical devices with intricate and complex structures that are delicate and require special methods and technical requirements for cleaning, disinfection, and sterilization^[12]^(e.g., endoscopic tools and micro-instruments for neurosurgery) ; ➁ Instruments that are reusable.

Exclusion criteria: ➀ instruments that need to be scrapped and ➁ instruments that have not reached the scrapping level but whose repair cost exceeds the value of the instrument itself.

### 2.4 Interventions

#### 2.4.1 Experimental Group Interventions

The LSS management method based on Quality Circle activities was applied, including five stages: Define, Measure, Analyze, Improve, and Control.

##### 2.4.1.1 Define Stage

1. Project Team Formation: The team consisted of 13 members, including one team leader, two coaches, two internal trainers, and eight team members. The team was characterized by its cross-departmental and multi-industry composition, combining logistics and clinical expertise with strong working capabilities and rich management experience.
2. Customer Needs Analysis: Surgeons, clinical and operating room nurses, and the finance department were identified as internal customers, while patients and higher-level supervisory personnel were identified as external customers. By analyzing the needs of internal and external customers, the main improvement directions were identified as improving the processing workflow of precision instruments and reducing the wear rate.
3. Project Definition: Deterioration of precision instruments was defined as any abnormality, malfunction, or damage that occurred during their use in the hospital. The wear rate of the precision instruments was calculated as (number of deteriorated instruments/total number of instruments in the same period) × 100%.
4. Project Scope: Using the Six Sigma macro process map (SIPOC model), which includes suppliers, inputs, processes, outputs, and customers, all decision points that might affect the process were identified. The process includes the use of instruments, pre-processing instruments, transporting instruments, sorting instruments, cleaning instruments, disinfecting instruments, drying instruments, inspecting and maintaining instruments, packaging instruments, sterilizing instruments, storing instruments, distributing instruments, and reusing instruments.
5. Project Plan and Progress Management: A Gantt chart was drawn, and the project was planned to be completed within 10 months.

##### 2.4.1.2 Measure Stage

1. Measurement Plan: Baseline data on instrument deterioration were collected through records of instrument repairs and scrappings from the medical engineering department and defect registration forms from the departments. The instruments were categorized into five types based on their degradation status: instruments to be scrapped, instruments with repair costs exceeding their value, instruments with repair costs lower than their value, instruments with degradation but no need for repair, and normal instruments.
2. Data Collection: According to the 5W2H method, the total number of precision instruments processed before the improvement from January 15 to April 30, 2023, was approximately 32,000, with 5,640 degraded instruments, resulting in a degradation rate of 17.63%. The wear rate was further categorized into ten distinct groups: instrument deformation, corrosion, missing parts, damage, lens wear, malfunction, aging, confusion, mismatch, and other forms of degradation. Through the Pareto chart, the focus of improvement was identified as instrument deformation, corrosion, and missing parts. The wear data were independently recorded by two trained staff members using standardized forms. Discrepancies were resolved through consensus with a third auditor from the Quality Management Office.
3. Objective Setting: Based on baseline data and interviews with 20 surgeons, 20 nurses, and 10 managers, combined with the hospital’s management capabilities, the target deterioration rate was set at less than 10%.

##### 2.4.1.3 Analyze Stage

1. Fishbone Diagram Analysis: A brainstorming session was conducted to analyze the causes of instrument deformation, corrosion, and missing parts from four perspectives: personnel, environment, materials, and methods. The fishbone diagrams were drawn accordingly. See Figure 1.
2. Cause and Effect Matrix Analysis: The causes identified through the fishbone diagram are arranged in rows and columns. Using the Cause and Effect Matrix matrix, the team collectively evaluated the relevance and degree of association between factors. Sixteen significant factors were selected based on the 80/20 principle, 16 significant factors were ultimately selected. See Figure 2.
3. Cause-and-Effect Diagram Analysis: Using an association diagram, complex relationships, such as cause-effect and means-end, were logically connected with arrows. The mean score was 31/16 (1.93). Factors with a negative absolute score of ≥2 were identified as root causes, resulting in the identification of seven root causes. See Figure 3.

**Figure 1.**
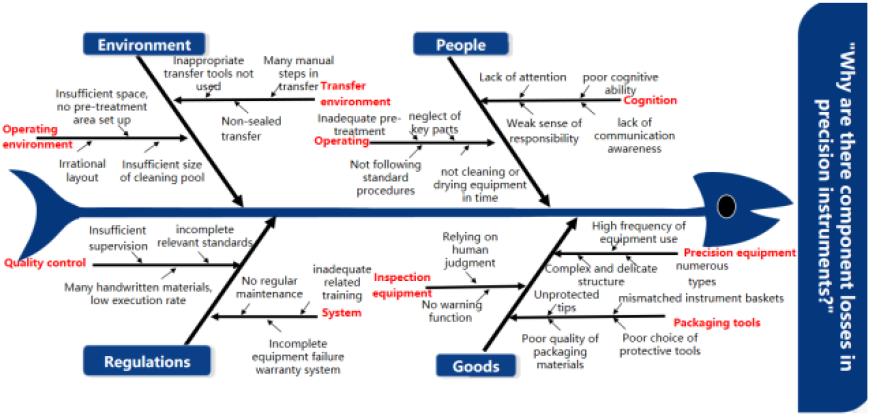
Fishbone Diagram Analysis

**Figure 2.**
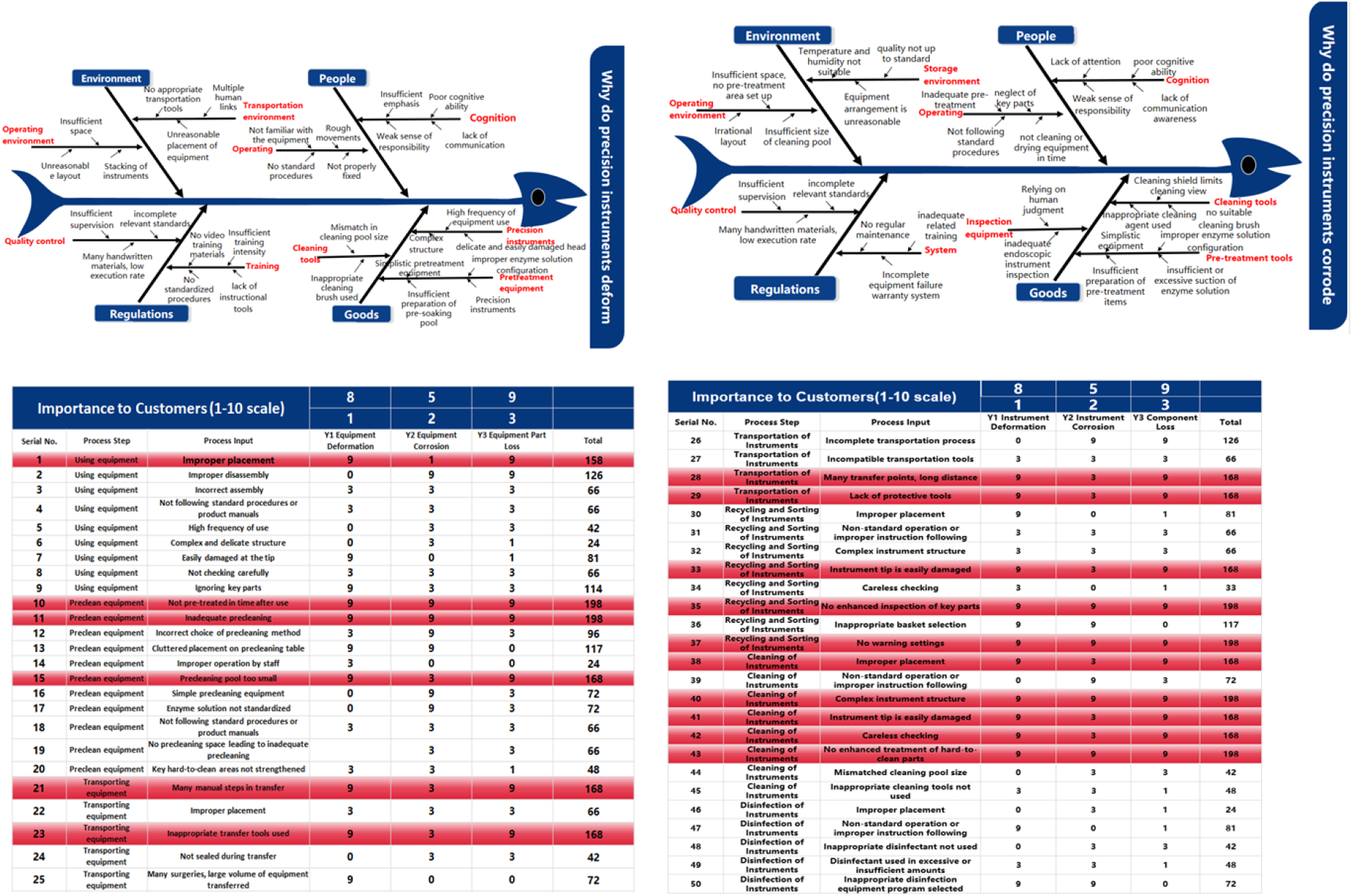
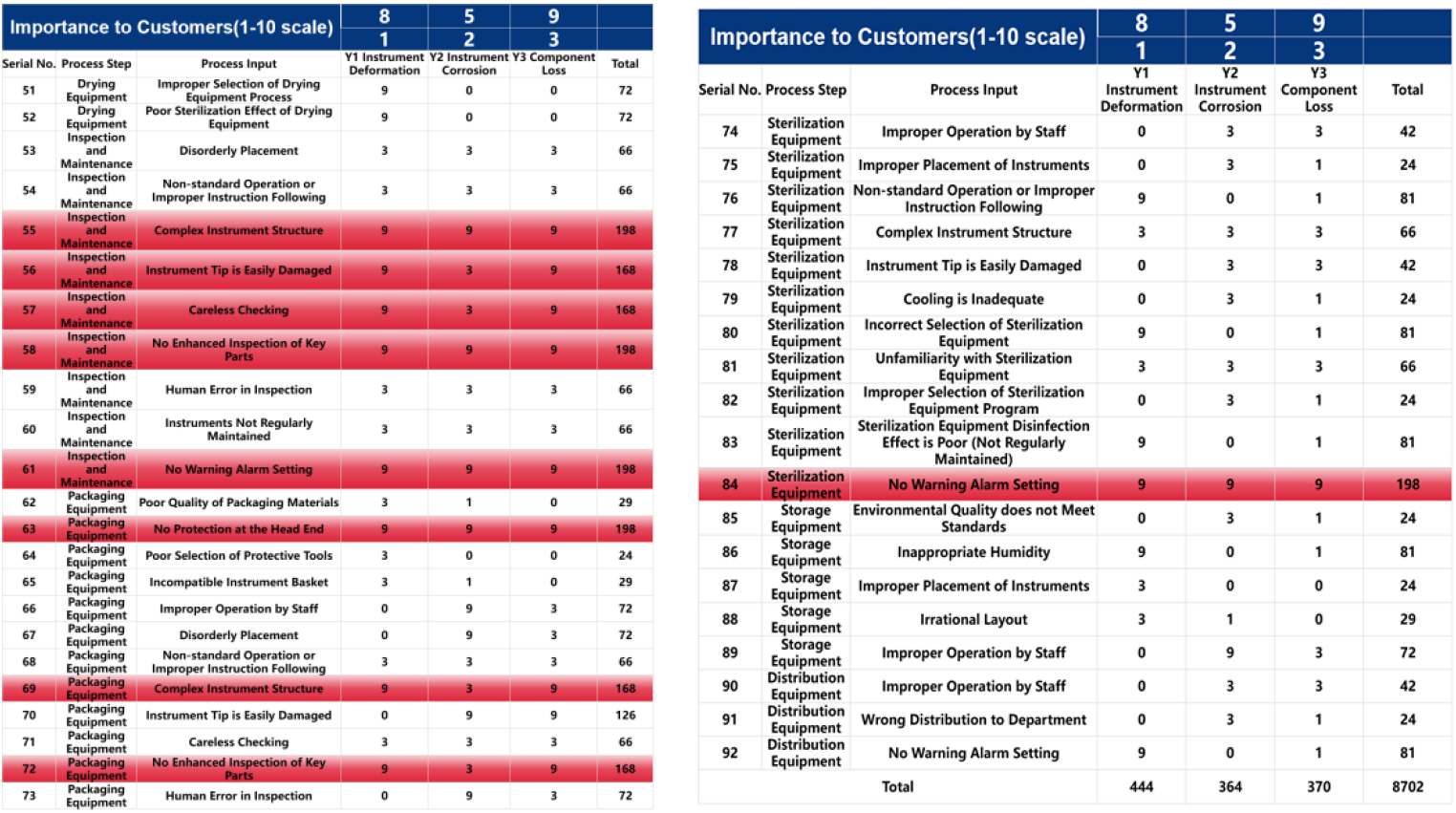
Cause-and-Effect (C&E) Matrix Analysis

**Figure 3.**
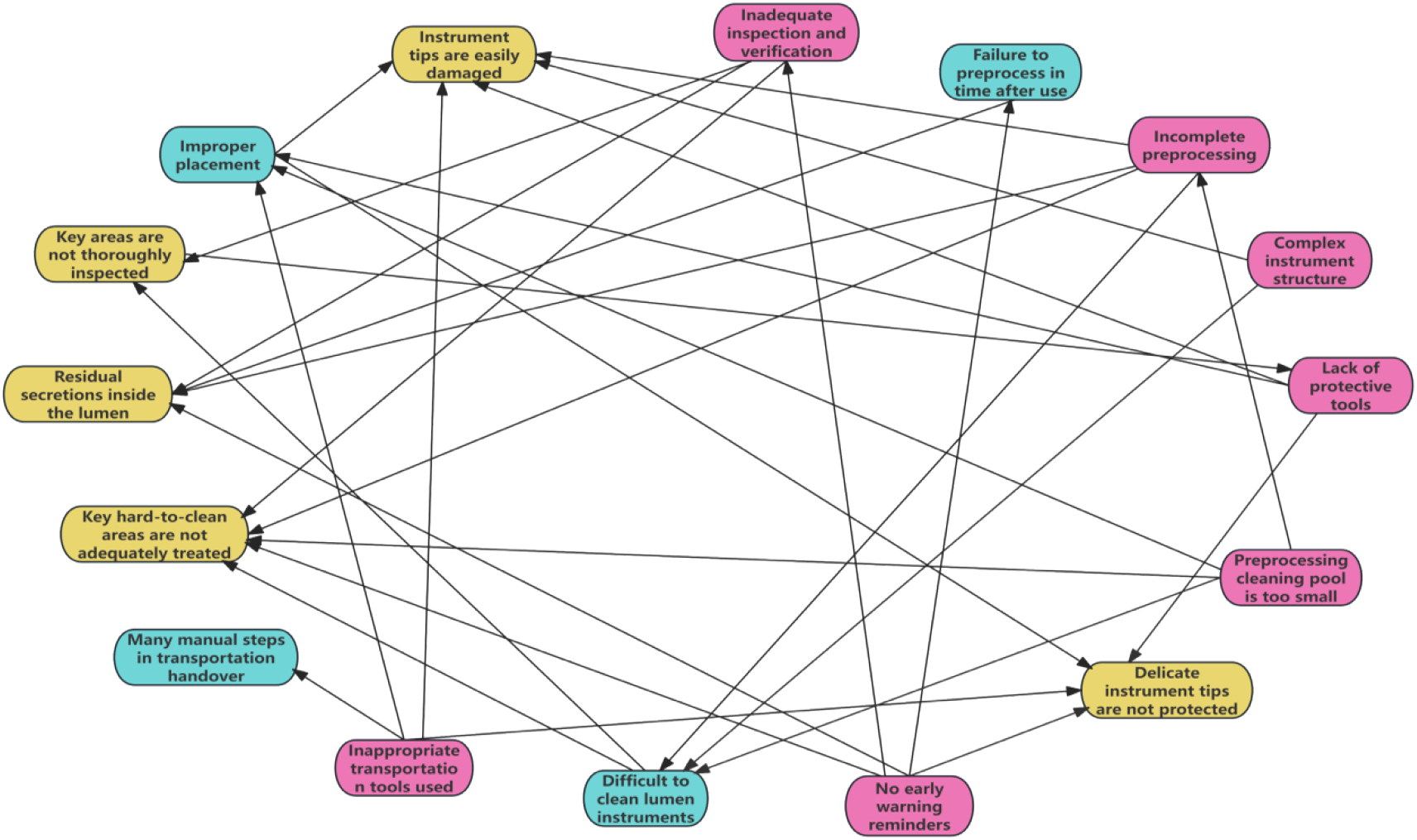
Cause-and-Effect Diagram Analysis

##### 2.4.1.4 Improve Stage

The team scored the feasibility, cost-effectiveness, and impact of the proposed solutions on a 1-5 scale (5 = excellent, 3 = acceptable, and 1 = poor). The total score is 165. Based on the 80/20 principle, solutions with scores greater than 132 were selected and merged into three major strategies.

1. Establishing an Intelligent Device Traceability and Sterile Supply Chain Quality Control Management System: An intelligent work platform was created through the management backend, the CSSD(Central Sterile Supply Department), and surgical/clinical functional islands. This system enables regional, multi-hospital, and hospital-wide management of precision instruments. Key processes and steps in the full lifecycle of precision instruments, from preprocessing to reuse, were linked with intelligent diagrams and early warning reminders. The “Eagle Eye” intelligent counting instrument was used for coding and identification of instruments, enabling patient, device, and instrument traceability. The system also facilitated visual management between the sterile supply center and multiple clinical departments, and implemented full-process quality control with specific assessment indicators.
2. Refining Transportation Management: Intelligent robots were used for transportation to reduce the manual handling processes. For high-value consumables, electric double-layer transport vehicles have been designed to prevent instrument compression, cross-contamination, and reduced transport time. For low-value consumables, transport vehicles of different colors and models were selected, and the peak number of instrument packs was set based on their size and weight.
3. Pre-processing Position Shift and Use of Portable Anti-Pollution Pre-processing Integrated Devices: The pre-processing position was moved forward to the operating room, returning surgical nurses to patient care and improving pre-processing effectiveness. A portable integrated anti-pollution pre-processing device was designed and used, featuring one-button operation and automatic solution preparation, effectively enhancing pre-processing quality and preventing instrument corrosion.

##### 2.4.1.5 Control Stage

1. The process map for precision instruments was revised to strengthen the supervision of key process steps and to ensure continuous quality improvement.
2. A quality control management process map for precision instruments was established to clarify management responsibilities, conduct ongoing supervision and inspection, calculate the wear rate regularly, and provide feedback to ensure continuous control of the project.
3. Standard operating procedures were developed for pre-processing, cleaning, and transportation of precision instruments, ensuring that the project was implemented in accordance with the established guidelines and could continue sustainably.

#### 2.4.2 Control Group Interventions

The use, transportation, and reprocessing of instruments were strictly implemented according to WS 310.2-2016 “Part 2 of the Hospital Sterile Supply Center: Specifications for Cleaning, Disinfection, and Sterilization Techniques’[^12^] and the instrument product instructions. The operating staff preprocessed the instruments immediately after use and transported them to the CSSD for cleaning, disinfection, and sterilization. Qualified instruments were transported back to the operating room or clinical departments for reuse, whereas unqualified instruments were reprocessed.

#### 2.4.3 Evaluation Indicators and Data Collection Methods

##### 2.4.3.1 Evaluation Indicators

1. Comparison of Wear Rates of pre-LSS and post-LSS
2. Direct economic benefits of the project.
3. Comparison of the satisfaction rates of surgeons and nurses before and after implementation of the LSS management method.

Satisfaction is divided into five levels (Very Satisfied, Satisfied, Neutral, Somewhat Dissatisfied, and Dissatisfied). Ratings were given for 10 questions across three major aspects: quality of equipment support, timeliness of equipment support, and service attitude of the staff. The Satisfaction Rate was calculated as follows: satisfaction rate = (Number of Very Satisfied + Number of Satisfied) / Total Number of Respondents × 100%.

##### 2.4.3.2 Statistical Methods

Statistical analyses were performed using SPSS 25.0 (IBM Corp., USA). Non-normally distributed data were analyzed using Mann-Whitney U tests, with effect sizes reported using Cohen’s d. Count data were described using sample size and percentage, and comparisons between groups were conducted using chi-square tests or rank-sum tests. For normally distributed measurement data, the mean ± standard deviation was used for the description, and independent sample t-tests were used for comparisons between groups. For non-normally distributed measurement data, the median and quartiles were used for the description, and rank-sum tests were used for comparisons between groups. The significance level was set at p =0.05. Normality tests (Shapiro-Wilk, p<0.05) indicated a non-normal distribution of deterioration rates; thus, Mann-Whitney U tests were applied to confirm significant differences (U=XX, p<0.001).

## 3 RESULTS

### 3.1 Comparison of Wear Rates pre-LSS and post-LSS

The wear rate of the precision surgical instruments in the experimental group was significantly lower (6.54%) than that in the control group (17.63 %) (P<0.001). The effect shift chart shows that by December 2024, the wear rate decreased to 6.09%, meeting the target. See Table 1 and Figure 4.

**Table 1:** Comparison of Wear Rates Between Groups [n (%)]

| Indicator | Experimental Group<br>(n=31,989) | Control Group<br>(n=32,000) | $\chi^2$ Value | P Value |
| --- | --- | --- | --- | --- |
| Deterioration Rate | 2,092 (6.54%) | 5,640 (17.63%) | 1850.456 | <0.001 |

**Figure 4.**
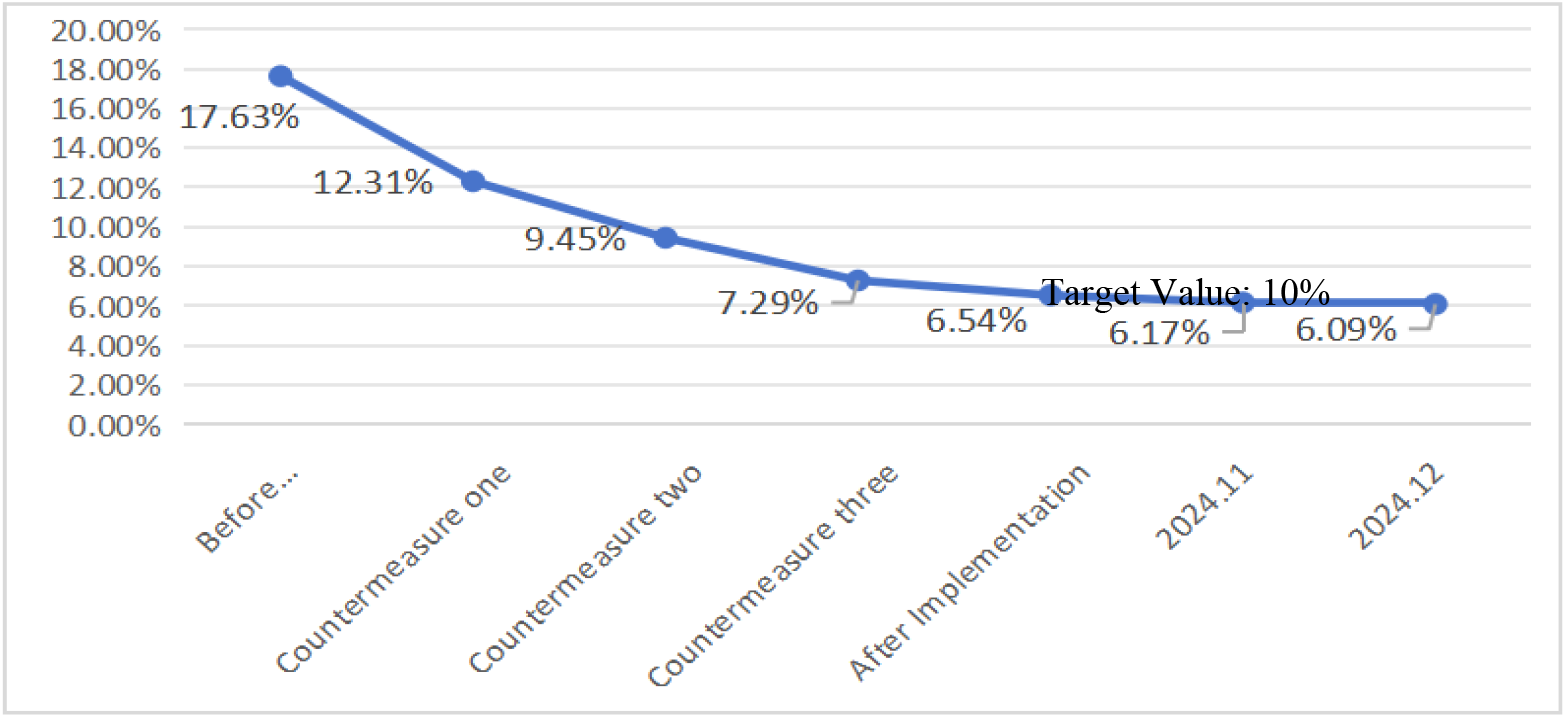
Effect Shift Chart of Wear Rate Reduction

### 3.2 Direct Economic Benefits of the Project

Based on data from the hospital’s medical engineering department on repair and scrapping costs, the cost savings from instrument repairs were calculated as 730,000 yuan (before improvement) to 310,000 yuan (after improvement) = 420,000 yuan. The cost savings from scrapping instruments ranged from 539,000 yuan (before improvement) to 190,000 yuan (after improvement) = 349,000 yuan. The total direct economic benefit is 769,000 yuan.

### 3.3 Comparison of Surgeon and Nurse Satisfaction pre-LSS and post-LSS

Surgeon satisfaction significantly improved from 83.33% (100/120) pre-intervention to 95.83% (115/120) post-intervention (P=0.002), with a Cronbach’ s α of 0.85 indicating high internal consistency.And nurse satisfaction increased from 86.67%to 98.33%(P=0.001). See Table 2.

**Table 2:** Comparison of Surgeon and Nurse Satisfaction pre-LSS and post-LSS [n (%)]

| Indicator | Pre-LSS | Post-LSS | $\chi^2$ Value | P Value |
| --- | --- | --- | --- | --- |
| Surgeon Satisfaction | 83.33% | 95.83% | 10.047 | 0.002 |
|  | ( 100/120 ) | ( 115/120 ) |  |  |
| Nurse Satisfaction | 86.67% | 98.33% | 11.772 | 0.001 |
|  | ( 104/120 ) | ( 118/120 ) |  |  |

## 4 DISCUSSION

The use of instruments in hospitals involves a series of processes, including use, preprocessing, transportation, sorting, cleaning, disinfection, drying, inspection and maintenance, packaging, sterilization, storage, distribution, and reuse by patients. Any issue in these processes can significantly impact the overall quality^[13]^. The Central Sterile Supply Department (CSSD) is a crucial department in hospitals for ensuring the hygiene and safety of medical instruments^[14]^. According to the health industry standards of the People’s Republic of China, reusable medical instruments should be returned to the CSSD for centralized management after use. The level of management directly affects the lifespan of the instruments and the effectiveness of clinical surgical treatments^[15]^. This study, conducted in collaboration with multiple hospital departments, applied the LSS management method to the entire lifecycle of the precision instruments. The results showed that the wear rate decreased from 17.63% to 6.54%, directly reducing the hospital medical costs by 769,000 yuan. In addition, the satisfaction of surgeons and nurses in the sterile supply department increased. This indicates that the LSS management method can promote improvements in CSSD management and enhance the management quality.

Depreciation constitutes the primary cost of the instruments. Higher utilization rates reduce per-procedure costs, whereas reduced deterioration extends instrument lifespan, enabling more frequent reuse^[16-17]^. Therefore, reducing instrument deterioration is significant in lowering instrument costs. The results of this study showed that the main types of precision instrument deterioration are deformation, corrosion, and missing parts. In the fishbone diagram, C&E matrix, and cause-and-effect diagram analyses, the primary reasons were identified as complex instrument structures, lack of protective tools, improper transportation tools, insufficient inspection and verification, lack of early warning reminders, inadequate pre-processing, and small pre-processing sinks. Based on these findings, improvements were made in preprocessing, transportation, and information management.

Pre-processing involves the removal of visible contaminants from instruments after use with optional humidification and rinsing. Instruments should be pre-processed as soon as possible at the point of use. Timely cleaning can reduce instrument corrosion caused by contact with blood, saline, or other chemicals (e.g., hemostatic agents) during surgery. However, instruments used in surgery are often covered with large amounts of blood, fat, mucus, and proteins, which can promote microbial growth and biofilm formation. Once dried, these contaminants are difficult to remove and can affect the subsequent disinfection or sterilization processes, leading to sterilization failure^[18-19]^. To prevent biofilm formation and microbial growth, staff in the operating room and sterilization departments should promptly remove large contaminants at the time of use. Therefore, instruments should be pre-processed immediately after use according to the manufacturer’s instructions and guidelines. In this study, the pre-processing position was moved forward to the operating room to address the issue of insufficient pre-processing owing to staffing shortages. A portable anti-pollution pre-processing integrated device was developed and used to effectively improve pre-processing quality and reduce instrument degradation. While the intelligent robot transportation system reduced manual handling, its scalability may be limited by upfront costs and staff training requirements, as observed in similar studies.

The transportation of instruments is an essential part of the reprocessing cycle of reusable instruments. Hospitals have different hardware facilities and transportation methods. Based on the issues identified in the current transportation process, this study implemented intelligent robot transportation to reduce the number of manual handling steps. For high-value consumables, electric double-layer transport vehicles have been designed to prevent instrument compression, cross-contamination, and reduced transport time. For low-value consumables, transport vehicles of different colors and models were selected, and the peak number of instrument packs was set based on their size and weight.

Several studies have shown that continuous monitoring and traceability programs in CSSD can effectively improve the cleaning, disinfection, and sterilization quality of instruments, ensuring professional, standardized, and standardized processes. This helps control hospital infections and ensure patient safety^[5,20]^. Based on clinical practice needs, this study established an intelligent device traceability and sterile supply chain quality control management system on an existing information reprocessing platform. The new system includes multidepartment collaborative management, intelligent diagram linkage, voice early warning reminders, intelligent counting, and full-process quality control. The results showed that the instrument degradation rate was significantly reduced and the average processing time for the instruments was shortened. This system is beneficial for improving the instrument management quality and ensuring patient safety in clinical settings.

## 5 STRENGTHS AND LIMITATIONS

This study involved collaboration between the hospital’s CSSD, operating room, information department, medical engineering department, and the quality management office. It introduced lean and six-sigma management to enhance safety awareness in precision instrument management, and focused on eliminating waste and creating value. However, this study has certain limitations. First, this single-center study was conducted at a tertiary teaching hospital with advanced resources. Future multicenter trials in community hospitals or low-resource settings are necessary to assess the generalizability of LSS interventions. Second, this study used a single quality management method to investigate instrument deterioration, which may have introduced some data biases. Future studies should combine longitudinal data tracking with qualitative interviews to explore staff’s perspectives on LSS implementation.

## 6 CONCLUSION

The full lifecycle management of precision instruments based on LSS can effectively reduce the deterioration rate, lower hospital costs, and improve the satisfaction of surgeons and nurses with the sterile supply department.This management model breaks through the traditional extensive management approach and actively explores the use of advanced management methods. It not only optimizes the management process of medical devices, but also enhances management efficiency. Its successful implementation can provide a reference for the quality management of medical devices in hospitals, and is conducive to promoting the overall improvement of hospital management levels, thereby better serving clinical medical work.

## Data Availability

Data cannot be shared publicly because of Unit Confidentiality Treaty. Data are available from contact XiaoXia Zhu for researchers who meet the criteria for access to confidential data.

## Acknowledgements

The authors extend their sincere gratitude to all anonymous reviewers, editors, and participants in this study, and the First Affiliated Hospital of Naval Medical University for their support.

## Author Contributions

XiaoXia Zhu and Jing Qin contributed to the original draft and methodology; Xia Zhou and HaiPing Chen collected and analyzed the data; and Rong Bao and CongBing Yan reviewed and edited the manuscript. All authors have reviewed the final manuscript.

### Declarations

Ethics approval and consent to participate: Not required, as the study involved medical devices.

Consent for publication: Obtained from the First Affiliated Hospital of Naval Medical University.

Competing interests: The authors declare no competing interests.

### Funding

This manuscript is supported by funding a granted from RongBao (Project No: 2023HXA01) and Jing Qin.(Project No: 2023HXA02).

